# Unmasking digital despair: A language model framework for predicting depression on social media

**DOI:** 10.64898/2026.09.21.26363552

**Authors:** Matthew Xie, Hangfeng He, Zidian Xie

## Abstract

Depression is one of the most prevalent mental health disorders globally. Social media content can reflect emotional states, offering real-time signals of depressive symptoms. This study aimed to develop a deep learning model to predict depression using Twitter/X data, and to characterize potential causes of depression using a large language model (LLM). Twitter/X posts from April 20, 2023, to July 24, 2024, were obtained using the key phrase (“I” or “me”) and “diagnosed with depression”. After data cleaning and GPT-4o labelling, 2,275 depressive Twitter users, and an additional 1,661 non-depressive users from tweets using the keyword “today”, were identified. A deep learning RoBERTa model for predicting depression was built on tweets from both user groups, with an 80/20 training–testing split. GPT-4o models were subsequently employed to analyze depressive users’ posts to understand potential causes for depression. The RoBERTa model achieved strong performance in predicting depression among Twitter users from their tweets, with an accuracy of 0.822, an F1 score of 0.855, and an AUC of 0.809. Common reasons for depression identified by GPT-4o models included societal pressure, low self-esteem, cultural influences, and identity-related challenges. These findings highlight the potential of deep learning models in early screening of depression using social media data. Insights into potential reasons for depression may inform targeted prevention strategies, public health interventions, and improved mental health support for at-risk populations.

## Introduction

Depression is one of the leading causes of disability worldwide, with its global presence increasing by nearly 50% over the past three decades (1). Lifetime recurrence rates are estimated to be as high as 90% in certain populations (1). Clinically, depression is characterized by persistent sadness, diminished interest or pleasure in activities, and disturbances in cognition, appetite, and sleep, all of which substantially impair daily functioning (2). When untreated, these symptoms can accumulate and intensify, contributing to severe outcomes, including suicidal behavior. Globally, depression is associated with approximately 700,000 suicide deaths each year (3), many of which may be preventable with timely access to evidence-based treatment and support services. According to the World Health Organization, depression arises from complex interactions among biological, psychological, and social determinants (3). Contributing risk factors include adverse life events (e.g., bereavement), chronic stress, harmful health behaviors such as alcohol misuse, and physical inactivity.

In addition to the devastating health effects, depression imposes a substantial economic burden with annual costs estimated to exceed $325 billion in the U.S. alone (1). Furthermore, depression is widespread in the U.S. with more than 16% of adults and 4.4% of children experiencing depression (2). Data from the 2020 Behavioral Risk Factor Surveillance System (BRFSS) indicate that depression prevalence in the U.S. is highest among young adults aged 18–24 years (21.5%), women (24.0%), non-Hispanic White individuals (21.9%), and those with less than a high school education (21.2%) (4). Globally, an estimated 280 million individuals are affected by depression (3). These substantial prevalence rates underscore the widespread burden of depression nationally and worldwide and highlight an urgent need for effective prevention and intervention strategies.

While advancements in existing depression treatments remain essential, ensuring adequate access to care for individuals at risk for, or already experiencing, depression is equally critical. Evidence from a study conducted in British Columbia indicates that only 53% of individuals with depression received at least minimal counseling/psychotherapy or minimally adequate antidepressant therapy (5). This substantial treatment gap underscores the need for effective strategies to identify individuals who require intervention.

Social media platforms provide a promising avenue for mental health detection, with use strongly associated with depression risk among U.S. young adults (6). A recent review highlights social media text as a valuable resource for predictive modeling of depressive symptoms (7) (8) (9). Specifically, studies have analyzed linguistic and behavioral cues in tweets prior to the reported onset of depression, noting that Twitter provides insights into psychosocial circumstances and potentially other reasons (10). Similarly, other computational studies identified language patterns across users with different mental health conditions (11) (12). One prior study used conventional machine learning methods (e.g., support vector machines) to detect depression on Twitter, achieving at best ∼70% accuracy (10). More recent natural language processing (NLP)-based models have reported accuracies of up to 88% on a public Kaggle dataset; however, depression was defined by keyword filtering, potentially biasing the ground-truth labels (13). In a study analyzing 9,300 twitter posts, relying on keyword queries alone are found to be insufficient for public health monitoring because of the post context changing the meaning of the keyword (14). More generally, critical reviews of computational mental health research have identified concerns regarding construct validity and data annotation, noting the lack of reflection or evaluation of a new data annotation technique across numerous studies (9).

In contrast to simpler NLP models, deep learning architectures such as Bidirectional Encoder Representations from Transformers (BERT) and the Robustly Optimized BERT Pretraining Approach (RoBERTa) have demonstrated substantially higher performance in social media– based depression detection, with accuracies up to 98% in Reddit studies (15). Similarly, a RoBERTa-based Twitter study achieved 90% accuracy with F1 scores of 0.86–0.96 across depression severity levels (16, 17). Although prior studies have demonstrated that social media posts before a diagnosis may contain information about depression risk (11) (18), many widely used datasets and classification approaches continue to rely on depression-related keywords, self-disclosures, or posts containing explicit mental-health terminology, limiting their application in early screening and prospective risk prediction based on naturally occurring conversation and discourses on social media.

Using Twitter (X) data, this study aims to examine depression-related content based on users’ self-reported diagnoses, leveraging a widely used platform (e.g., Twitter/X) for large-scale mental health research (19). This study aims to build a RoBERTa predictive model for depression based on naturally occurring Twitter posts that do not explicitly reference depression, and more importantly, use large language models (GPT-4o) to further understand potential reasons for depression shared on social media.

## Materials and Methods

### Ethics Statement

This research did not require institutional review board oversight because it analyzed fully anonymized, publicly accessible social media records without any researcher interaction or private data access. All data were deidentified prior to analysis to maintain user privacy.

### Data Collection

Twitter data was obtained through the Meltwater platform, a commercial social media monitoring and analytics service that enables the systematic collection of large-scale social media content for research purposes. To identify posts related to self-reported depression diagnosis, we retrieved all English-language tweets containing the phrase “I” or “me” or “mine” in combination with “diagnosed with depression” using case-insensitive matching. This approach has been widely adopted in prior research to identify users self-reporting a depression diagnosis (20, 21). Between April 20, 2023, and July 24, 2024, a total of 18,321 tweets from 17,215 unique Twitter users were identified. For the control dataset of users who did not claim a depression diagnosis, we sought to identify a keyword that was predominantly neutral in sentiment and had been used in comparable studies (22), thereby minimizing the potential for systematic bias in either direction. For this purpose, we collected 20,000 English-language tweets posted between January 1, 2024, and July 31, 2024, containing the keyword “today” using case-insensitive matching.

### Data Cleaning

We first cleaned the depression dataset by further filtering the depression status of all potentially depressed users. Because our objective was to identify user self-reported depression, we removed posts containing the phrase “not diagnosed with depression,” which also matched our keyword criteria. We also excluded all retweets as reposted content does not necessarily reflect the user’s own experiences or mental health status. Following these exclusions, 5,562 posts from 5,027 unique users remained in the depression dataset. For the control dataset, we removed all users whose tweets contained mental health–related terminology, including “depression”, “depressed”, “depress”, “failure”, “hopeless”, “nervous”, “restless”, “tired”, “worthless”, “unrested”, “fatigue”, “irritable”, “stress”, “dysthymia”, “anxiety”, “adhd”, “loneliness”, “lonely”, “alone”, “boredom”, “boring”, “fear”, “worry”, “anger”, “confusion”, “insomnia”, and “distress”, to minimize the likelihood of unintentionally including users who might be potentially depressed (23, 24). After filtering, 19,490 posts from 19,321 unique users remained in the control dataset.

### Validation of Depressed Users

To further validate Twitter users labelled with depression, we randomly sampled 316 tweets (302 unique users) from the depression cohort for manual annotation. We developed two nested classification labels to characterize depression status: “ever depressed?” and “still depressed?”.

The “ever depressed” variable will be determined based on if the Twitter/X user had received a depression diagnosis at some point in their lifetime. Among those tweets labelled with “ever depressed”, they were further classified as “still depressed” if the Twitter/X user was currently experiencing depression. Manual annotation was conducted independently by two trained human coders. Discrepancies were discussed, and a consensus labeling protocol was established and applied across all sampled tweets. This iterative refinement continued until interrater reliability exceeded Cohen’s kappa of 0.8.

The human-annotated 316 sampled tweets served as the ground truth to optimize prompts for the large language model GPT-4o via an Application Programming Interface (API) until the GPT-4o model achieved an F1 score of 0.91, indicating strong model performance. Then, optimized prompts for GPT-4o were used to label the remaining depression dataset. The optimized prompts was provided with explicit instructions regarding label definitions and instructed to account for tweets containing quoted text (Table S1).

### Collection of Additional Tweets for Identified Twitter Users

For the identified and validated 2,464 unique users classified as “still depressed”, we retrieved all tweets posted by these users between August 3, 2023, and August 5, 2024, from the Meltwater database. To construct a comparable control cohort, we randomly sampled 2,500 unique control users (not depressed) and collected their tweets over the same period.

We removed all retweets and all tweets containing the phrase “diagnosed with depression” from tweets authored by users in the depression cohort. Additionally, words such as “depress,” “depression,” or “depressed” were excluded from all tweets to prevent the predictive models from relying on explicit depression-related terminology as a marker for classification. To maximize both the number of users and the total tweet volume, we randomly sampled up to 50 tweets per user from those with more than 50 tweets in our dataset. Twitter users with fewer than 50 available tweets were excluded from this stage to maintain consistent representation across users. The final depression dataset included 2,275 Twitter users, each with 50 tweets.

For the control dataset, we retrieved posts for all users in a manner similar to that of the depression dataset. We removed any user with tweets containing the phrase “diagnosed with depression” to minimize the likelihood of misclassification within the control group. Similarly, we randomly sampled up to 50 tweets per control user with more than 50 tweets and excluded users with fewer than 50 tweets. The final control dataset included 1,661 Twitter users with no self-reported depression, each with 50 tweets. A summary of all the preprocessing steps is shown in Fig S1.

### RoBERTa Predictive Model for Depression

We employed the Robustly Optimized BERT Pretraining Approach (RoBERTa) model, a transformer-based deep learning architecture widely used for natural language understanding tasks. Compared with the original BERT framework, RoBERTa is pre-trained with larger batch sizes, longer training durations, dynamic masking strategies, and optimized hyperparameters, resulting in substantially improved performance across multiple benchmark datasets (25).

RoBERTa processes text by tokenizing input sequences and learning contextualized word representations via multi-layer self-attention. Its expanded training corpus and enhanced vocabulary coverage offer advantages over BERT for analyzing large-scale social media text data (26). Prior work has demonstrated that RoBERTa outperforms traditional machine learning approaches for depression detection in social media data (15, 27). In the present study, we fine-tuned the RoBERTa model using the AdamW optimizer. The final dataset containing tweets from 2,275 users with self-reported depression and 1,661 users with no self-reported depression was divided into training and testing datasets using an 80:20 split, which was used to train and test the RoBERTa model. Model performance was assessed on the test dataset using accuracy, AUC, and F1 scores.

### Identification of Potential Reasons for Depression

GPT-4o is a state-of-the-art large language model with advanced natural language understanding and text analysis capabilities, demonstrating substantially improved performance compared with earlier GPT versions (28). Its ability to process large volumes of text efficiently makes it well-suited for analyzing the extensive Twitter datasets used in this study. We used GPT-4o to identify potential reasons for depression among users classified as depressed. For each depressed user, we extracted up to 50 of their most recent tweets posted on or before their anchor tweet (i.e., the tweet in which they self-reported a depression diagnosis). For users with fewer than 50 tweets, all available pre–anchor tweets were included to maximize data retention. This resulted in a total of 2,453 depressed users included for the reason identification phase of the study. GPT-4o was then applied using a structured prompt to assign one potential reason for depression to each user based on their historical tweets (Table S1). For the first round, we asked the model to produce numerous highly specific reasons that might be related to each other. To improve interpretability and reduce redundancy, the ChatGPT model was subsequently used to consolidate these reasons into 16 broader thematic categories, which were then used to re-label potential reasons for depression using the GPT-4o model with appropriate prompts (Table S1). A single most representative category was assigned to each depressed user.

## Results

### Depressed Twitter User Prediction Using the RoBERTa Model

Based on 50 tweets for Twitter users who either self-reported a depression diagnosis (2,275 users) or had no self-reported depression diagnosis (1,661 users), the RoBERTa deep learning model demonstrated strong model performance in predicting depressed Twitter users, achieving an AUC score of 0.809, an accuracy of 0.822, and an F1 score of 0.855.

### Potential Reasons for Depression

By analyzing tweets posted by 2,453 users who self-reported a depression diagnosis using GPT models, we identified more than 500 distinct reasons initially, which were further grouped into 16 categories. Table 1 provides detailed information about the 16 categories, with emotional and psychological challenges encompassing issues such as struggles with self-confidence and emotional instability, and health and well-being capturing the psychological impact of physical health conditions. Among the 2,453 users identified as depressed, 1,402 reported specific reasons for their depression, whereas the remaining 1,051 did not provide any reasons. As shown in Table 1, the most common reasons for depression among depressed Twitter users were emotional and psychological challenges (18.34%, 450/2,453), health and well-being-related concerns (9.74%, 239/2,453), and trauma and abuse (7.58%, 186/2,453).

**Table 1.** Potential Reason Categories for Depression.

| Reason Category | Frequency (%)<br>n=2,453 | Description |
| --- | --- | --- |
| **Emotional and Psychological Challenges** | 450 (18.34%) | Covers self-doubt, self-worth struggles, overthinking, paranoia, anxiety, emotional instability, mood swings, guilt, shame, loneliness, existential dread, self-sabotage, and pretending happiness. |
| **Health and Well-being** | 239 (9.73%) | Encompasses chronic pain, illness, disability, physical exhaustion, medication side effects, reproductive health concerns, medical misdiagnosis, access to healthcare, and the psychological impact of physical health struggles. |
| **Trauma and Abuse** | 186 (7.58%) | Encompasses childhood trauma, past trauma, physical abuse, emotional abuse, sexual abuse, domestic violence, parental neglect, and unresolved grief from loss. |
| **Relationships & Social Challenges** | 138 (5.63%) | Includes romantic breakups, infidelity, trust issues, misunderstandings, social rejection, parental conflicts, family |
|  |  | conflicts, friendship loss, abandonment, and societal expectations in relationships. |
| **Work and Academic Stress** | 82 (3.34%) | Includes academic stress, job loss, financial instability, workplace stress, overwork, burnout, career uncertainty, economic hardship, and struggles with balancing responsibilities. |
| **Self-Harm, Addiction, & Maladaptive Coping** | 69 (2.81%) | Covers self-harm, substance use, compulsive behaviors, eating disorders, gambling, overspending, and suicidal thoughts. |
| **Body Image and Appearance Concerns** | 56 (2.28%) | Includes body dissatisfaction, weight gain, dieting struggles, societal beauty standards, fitness struggles, and pressure to look a certain way. |
| **Social Isolation and Lack of Support** | 53 (2.16%) | Covers loneliness, social rejection, feeling misunderstood, lacking a support system, feeling detached from others, and struggling with social anxiety. |
| **Self-Identity and Personal Growth** | 38 (1.55%) | Covers gender identity struggles, self-expression concerns, cultural identity conflicts, discrimination, societal beauty standards, body image dissatisfaction, self-acceptance, and imposter syndrome. |
| **Loss and Grief** | 32 (1.30%) | Includes grieving a loved one, loss of friendships, childhood loss, mourning past experiences, adjusting to major life changes, nostalgia, and feelings of life moving too fast. |
| **Societal and Cultural Pressures** | 19 (0.77%) | Includes political frustration, discrimination, racism, misogyny, unrealistic societal expectations, gender roles, oppression, and pressures related to tradition or faith. |
| **Financial Hardships** | 16 (0.65%) | Covers poverty, financial instability, debt, difficulty affording healthcare, economic inequality, and job insecurity. |
| **Safety, Violence, and Harassment** | 12 (0.49%) | Encompasses harassment, physical violence, online threats, unsafe living conditions, political violence, war trauma, and domestic violence. |
| **Parenting and Family Responsibilities** | 5 (0.20%) | Covers parental pressure, parenting stress, pregnancy concerns, family burdens, caregiving, childhood neglect, and generational conflicts. |
| **Mental and Physical Exhaustion** | 4 (0.16%) | Covers burnout, overworking, sleep deprivation, sensory overload, life responsibilities, emotional exhaustion, and difficulty maintaining balance. |
| **Life Purpose and Existential Struggles** | 3 (0.12%) | Includes lack of direction, feeling unfulfilled, questioning personal purpose, career regret, and feeling “stuck” in life. |
| **None** | 1,051 (42.85%) | No clear reason. |

## Discussion

Based on tweets posted by Twitter users who self-reported a depression diagnosis, we have successfully built a state-of-the-art deep learning model (RoBERTa) for predicting depressed Twitter users, which offers a robust predictive model for identifying potential depression among Twitter/X users based on their shared posts online. Furthermore, we have characterized potential reasons for depression shared by those users. The most frequently identified reasons for depression were emotional and psychological challenges, followed by trauma and abuse, as well as health and well-being–related concerns. Collectively, these findings highlight the potential of integrating deep learning models and large language models for scalable depression detection and understanding using social media data. This approach may support future efforts in early risk identification and inform population-level prevention and intervention strategies for depression.

Twitter data have been widely used for depression detection, as social media platforms increasingly serve as outlets for individuals to express emotions and document aspects of their daily lives (16). Based on Twitter posts, we developed a predictive RoBERTa model that effectively identified users who are depressed. A recent review by Ta et al. reported that long short-term memory (LSTM) and convolutional neural network (CNN) architectures have historically been the most commonly used deep learning approaches for depression detection on social media (29). In contrast, the application of transformer-based models such as RoBERTa represents a more recent methodological advancement and yielded strong predictive results in our study. Prior work has also shown the effectiveness of RoBERTa in related contexts. For example, Zaman et al. applied the RoBERTa model to Twitter data collected between 2009 and 2016 to predict multilevel depression severity, achieving an accuracy of approximately 90% (27). In the study by Zaman et al., Twitter data used for training RoBERTa model contained tweets mentioning depression or related terms, which might limit its application for early screening and detection of users who have high risk of depression but did not mention depression or related symptoms in their social media posts. In contrast, in our study, we removed words containing the root word “depress” from all tweets prior to training the model, which potentially eliminates the possibility that the predictive model partly, if not completely, relies on these words to predict depression. Therefore, our findings extend this literature by using naturally occurring social media posts without mentioning depression to predict the risk of depression, which broadens its application for early detection. Additionally, while the neural network–based system SenseMood achieved high performance for depression detection on social media, it relied on multimodal data, including images, which are not consistently available to all users and may limit scalability (30). In contrast, the text-based approach employed in this study enables broader applicability across users who primarily engage through textual content, which is more accessible.

This study leverages recent advances in large language models (LLMs) to enhance understanding of depression on Twitter by characterizing potential underlying, user-level reasons for depression—an aspect that has received limited attention in prior social media–based depression studies. Our study identified the most frequently mentioned reasons for depression on Twitter— emotional and psychological challenges, trauma and abuse, and health-related concerns—align closely with established clinical frameworks described by the National Health Service and the Cleveland Clinic (2, 31). The predominance of emotional and psychological challenges may reflect a greater willingness among users to publicly express internal emotional states on social media compared with more sensitive experiences such as trauma or physical illness.

Characterizing the relative prevalence of self-reported reasons for depression may support population-level mental health surveillance, which provides us the valuable guidance on resource allocation and targeted outreach.

While this study successfully developed a predictive model for depression detection on Twitter, several limitations should be acknowledged. First, the overall sample size was relatively modest, and some users had a limited number of available posts, which might limit model performance. Thus, increasing the number of users and posts in future studies may further enhance model robustness and predictive performance. Second, identification of depression was mainly based on self-reported depression diagnosis, which could introduce some biases without clinical validation. Third, the lack of demographic information on Twitter users limited our ability to characterize the demographic distribution of users with self-reported depression. Another limitation is the reliance on manual annotation and GPT-4o-based annotation for labeling users as depressed or non-depressed. Although the interrater agreement and GPT-4o model performance were high in this study, both modes of annotation are inherently subject to error and human annotation risks interpretive variability, which may introduce some degree of label noise and affect dataset validity. Additionally, the characterization of potential reasons for depression in this study relied primarily on large language models (e.g., GPT-4o). While these models are powerful tool in understanding and analyzing large-scale text data, they may subject to bias and hallucination. In future studies, analyses should be conducted under the guidance of mental health professionals, with iterative human review to ensure clinical validity and interpretive accuracy. Finally, the RoBERTa model was trained exclusively on Twitter data, limiting its generalizability to other social media platforms. Linguistic patterns and user behaviors may differ substantially across platforms. Future work could address this limitation by extending model training and validation to datasets from other platforms, such as Reddit or Instagram, as data becomes accessible.

## Conclusion

By leveraging large-scale textual data from both depressed and non-depressed Twitter users, this study successfully developed a RoBERTa deep learning model capable of predicting depression on social media. In parallel, large language models were successfully applied to characterize prominent, user-level reasons that might be associated with depression. Together, these approaches demonstrate the potential of integrating transformer-based deep learning models and large language models to enhance the understanding of depression on social media platforms.

The proposed framework provides a scalable foundation for future research in digital mental health surveillance and may inform the development of data-driven strategies aimed at early detection and prevention of depression.

## Acknowledgements

We thank the help from Zeliang Zhang for training the RoBERTa model. The authors declare no potential conflicts of interest with respect to the research, authorship, or publication of this article. The authors received no external financial support for the research, authorship, or publication of this article. No copyrighted material, surveys, instruments, or tools were used in the research described in this article.

## Contributions

MX: Conceptualization, Data curation, Formal analysis, Investigation, Methodology, Project administration, Validation, Visualization, Writing-original draft, Writing-review & editing. HH: Supervision, Methodology, Writing-review & editing. ZX: Conceptualization, Data curation, Project administration, Supervision, Writing-review & editing.

## Data Availability

The Twitter data are publicly available from the Twitter/X website: https://twitter.com/home?lang=en.

## Supporting information

**Fig S1. Flowchart of Twitter data preprocessing**.

**Table S1. Prompts for GPT models**.

